# State Adoption of Paid Sick Leave and Cardiovascular Disease Mortality Among Adults Aged 15-64 in the United States, 2008 to 2019

**DOI:** 10.1101/2023.12.05.23299537

**Authors:** Sam Swift, Lexi O’Donnell, Brady Horn, Katrina Kezios, Tali Elfassy, Julie Reagan, Adina Zeki Al Hazzouri, Tracie Collins

**Author notes:** **Corresponding Author Information:** Samuel Swift, PhD, MPH, Assistant Professor, College of Population Health, University of New Mexico Health Sciences Center.

## Abstract

**Background:** Cardiovascular Disease (CVD) is the leading cause of death in the United States and may be prevented through improved working conditions. The United States is one of the few high-income nations that does not guarantee paid sick leave (PSL) at the federal level. Our objective was to examine the relationship between state-level PSL policies and CVD mortality.

**Methods:** We used quasi-experimental event study methods to examine the relationship between implementing a mandatory PSL policy for all employees at the state-level and county-level CVD mortality rates using National Center for Health Statistics data among working-age adults aged 15 to 64 over time from 2008 to 2019. During this time, 11 states implemented PSL policies. We examined the annual CVD mortality rates (2008-2019) in 1054 counties from all 50 states and Washington D.C., accounting for approximately 88% of the United States population in this analysis.

**Results:** We found that in the Northeastern region of the United States, there were drops in the CVD mortality rate for persons ages 15-64 for all years after PSL was implemented, ranging from 7.1 fewer deaths per 100,000 persons (β=-7.1, 95% CI = -9.7, -4.4) seven years post-treatment to 2.7 fewer deaths two years post-treatment (β=-2.7, 95% CI= -5.1, -0.3).

**Conclusion:** Our results support the use of state-level PSL policies to reduce county-level CVD mortality rates, especially in the Northeastern United States.

## INTRODUCTION

Cardiovascular disease (CVD) is the leading cause of death in the United States (US)^1^. There are established and growing socioeconomic disparities in CVD, with low-income persons and persons experiencing unemployment at a higher risk of experiencing as well as pre-maturely dying from CVD^2^. While CVD mortality in the US has declined in the past 50 years, the rate of decline may be slowing^3^,and disparities between high and low-income groups are persistent for CVD outcomes^4, 5^.

Working years are a key time in the life course for the accumulation of stress and risky behaviors that can be precursors for cardiovascular disease^6^. In addition to a well-established literature on individual-level factors and CVD, there is growing evidence suggesting the importance of policies designed to improve working conditions, such as Paid Sick Leave (PSL), as tools for eliminating racial and socioeconomic disparities in health in the US^7, 8^. Long hours^9^, psychosocial job strain^10^, and access to PSL^11^ are all aspects of employment conditions that are associated with health, including measures of CVD. For example, in a US cohort, working over 46 hours in a week was observed to be associated with self-reported CVD^9^, and another European study estimated between 8.8% and 10.4% of CVD may be attributable to job strain^12^. Other research found that precarious employment (characterized by a lack of contractual agreements, low pay, temporary status, and lack of collective bargaining) was associated with self-reported CVD^13^. Additionally, having PSL through one’s job was associated with a 24% reduction in the risk of dying from heart diseases, in a longitudinal sample of American adults aged 18-85^11^. While most prior research either evaluated individual-level factors or individual workplace PSL, there is currently limited research on state-level PSL policies and CVD trends and outcomes. Given the findings on individual workplace PSL, it is important to evaluate the potential of state level polices as tools for CVD prevention.

Guaranteed PSL for all workers can influence CVD outcomes by improving working conditions and consequently promoting healthy behaviors. Yet, the United States is one of few high-income nations that does not guarantee PSL at the federal level^14^, and is one of the very small number of high income countries that does not have some policy at the federal (or equivalent) level securing this benefit for most workers in the country^8^. However, since 2012, several US states have instituted state-level PSL policies^15^. This provides an opportunity for a quasi-experimental study comparing CVD mortality in PSL policy-implementing states to states that did not implement these policies. Thus, the objective of this study was to assess the relationship between the implementation of state-level PSL and CVD mortality among working-age adults. We hypothesized that the implementation of PSL would lower CVD mortality in implementing states vs. those that didn’t implement PSL.

## METHODS

### Study Population

The unit of analysis for this research is the county. We used data from the Centers for Disease Control Wide-ranging Online Data for Epidemiologic Research (CDC WONDER) database and CDC WONDER Underlying Cause of Death 1999-2020 database^16^, an online dataset of mortality data derived from the death certificates of all US residents. While CDC WONDER may include data from all United States counties and Washington, D.C., CDC WONDER suppresses data based on fewer than 20 deaths within a year, so these analyses did not include data from sparsely populated or wholly uninhabited counties. This method is used to protect the confidentiality of decedents and also ensures that we only use stable rates^17^. We restricted county level mortality rates to working age persons age 15-64. We chose this range as it is the range available from CDC WONDER that is the closest approximation of the working age in the US, where people can begin work at a minimum of 14 years^18^, and most Americans retire around age 65^19^.

As such, our population included all U.S. counties with stable death rates as described above. We set our baseline to year 2008, which is four years before the first state implemented a PSL policy. We ended our panel in 2019 to avoid any complexities introduced by the 2020 COVID-19 pandemic.

### Exposure: State-level Paid Sick Leave Policy

We reviewed state PSL policies using the National Conference of State Legislatures website^20^ and a recent academic review of PSL policies in the US^15^, taking data on policies from the same time period as our panel (2008-2019). We included the policies of the 50 US states and the District of Columbia. We operationalized these policies as a binary (yes/no) exposure, with the intervention beginning in the year a state policy was implemented. The states (and Washington D.C.) that implemented these policies during this timeframe were Connecticut (2012), Washington D.C. (2014), California (2015), Massachusetts (2015), Oregon (2016), Arizona (2017), Vermont (2017), Maryland (2018), Rhode Island (2018), New Jersey (2018), and Washington (2018). While Michigan also implemented this policy in 2019^15^, we did not include this state as there would not be post-policy time in our panel for this state. No states repealed these policies during our panel.

### Outcome: Cardiovascular Disease Mortality

To focus on working persons most likely to be helped by this policy change, our primary outcome of interest was the county-level crude CVD mortality rate for persons ages 15 to 64. We defined CVD mortality using International Classification of Diseases codes for CVD, which include deaths from diseases of the heart (ICD-10 codes I00-I09, I11, I13, I20-I151), essential hypertension and hypertensive renal disease (I10, I12, I15) and cerebrovascular diseases (I60-I169)^21^. The CDC WONDER database uses deaths as defined above as the numerator and county-level population data provided by Surveillance, Epidemiology, and End Results Program for these rates^16^.

### Covariates

To control for time-varying confounding, we considered the following time-varying variables (from 2008 to 2019) in our models. These county-level covariates came from several different datasets. Annual county-level median income (in dollars) and percentage of a county living in poverty were accessed from the United States Census Small Area Income and Poverty Estimates (SAIPE). Both of these covariates were modeled by the United States Census using data from the American Community Survey, tax data, and other sources^22^. SAIPE defines poverty as family income falling below the federal poverty threshold for a given year^22^. We included annual county-level unemployment rates obtained from the Bureau of Labor Statistics^23^. We also included the annual percentage of a county without health insurance, obtained from the United States Census Small Area Health Insurance Estimates (SAHIE) database, which uses information from the American Community Survey and other sources to model an estimation of health insurance coverage by county^24^.

### Statistical Analysis

We employed quasi-experimental event study models to assess the effect of state-level PSL policies on CVD mortality. The advantage of these models is that with a disease like CVD, the trends in post-intervention effect of PSL may vary (the intervention may not be immediately effective), and the event study models allows for post-treatment trends over time. First, we mapped the year of implementation of each policy using the mapping function in Microsoft Excel^25^. For our primary analysis, we compared county-level CVD mortality among counties in states that implemented these policies to county-level CVD mortality among counties in states within the same United States Census Region (Northeast, South, West) that did not implement these policies to ensure that our control counties are similar to our treated counties. To group states, we used the United States Census Regions, which are Northeast, South, Midwest, and West^26^. To visualize trends in CVD mortality in treated and untreated states, we graphed the CVD mortality rate over time for each of the states that implemented these policies, compared to all non-PSL implementing states within the same region as the implementing states, displaying data from 2008 to 2019. Next, we display differences in average county-level baseline (2008) characteristics and changes in time-variant covariates (2008-2019) between our treated counties (within a PSL implementing state) and untreated counties (not within a PSL implementing state). Here, we tested for differences in means of these covariates between groups using independent samples t-tests.

A fundamental assumption of our event study models is that before treatment, the trends in outcome among the treated and untreated groups are parallel. To provide evidence that the two groups are comparable in terms of trend during the pretreatment period and assess differences in the crude age 15-64 CVD mortality rate post PSL implementation, we implemented an event study model, which takes the following form:

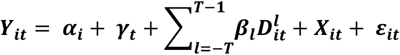

In this model, α_i_ and γ_t_ are fixed effects for counties and years, respectively, which is a common approach for policies that are implemented at different times in different places. This method is a subcategory of difference-in-difference models that use a two-way fixed effects estimator, which is a class or model that refer to the unit (county) and time (year-specific fixed effects that are included in this model. This event study model additionally includes lag (post-treatment) and lead (pretreatment) dummy variables for all counties in treated and untreated states, which is represented by 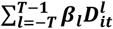.

The intuition of this model is that if the pretreatment dummy coefficients are null, then there is more evidence that the trends in outcome were parallel pretreatment and the post-treatment effects are closer to a causal effect of treatment, given that all other assumptions of these models are met. X_it_ is a matrix of the time-variant covariates mentioned above, and ε_it_ is the error term. We clustered standard errors at the level of treatment (state-level) for all event study models and weighted the models by the total population of each county.

Our main analysis implemented these models by comparing counties in treated states to counties in similar untreated states (states within the same census region). Due to the low implementation of this policy in the South (only one state and Washington, DC) and the lack of implementation of these policies in the Midwest, in our main event study models, we only made these comparisons within the Northeast and West regions. For this analysis, the Western region included the adopting states of Arizona, California, Oregon, and Washington and the non-adopting states of Colorado, Idaho, New Mexico, Montana, Utah, Nevada, Wyoming, Alaska, and Hawaii. The Northeastern Region included the adopting states of Connecticut, Massachusetts, Rhode Island, Vermont, and New Jersey and the non-adopting states of Maine, New Hampshire, New York, and Pennsylvania.

We conducted three sensitivity analyses. To assess effect modification by sex, we estimated event study models stratified by male and female sex (recorded as binary male/female in the CDC WONDER data) using the same time-varying controls, restricting comparisons to the same regions. Next, we estimated event study models using age-adjusted CVD mortality rates (age-adjusted but still limited to deaths among persons 15-64) as the outcome to see whether age adjustment changed the results. Finally, we estimated an event study model comparing counties within treated states nationwide to counties within untreated states nationwide.

All analyses were conducted in Microsoft Excel^25^ and R studio^27^. The data are publicly available from CDC WONDER for this analysis. The data and code can be viewed in a GitHub repository, as mentioned in the supplemental material.

## RESULTS

Of the 3,143 counties or equivalent geographic areas in the US, we conducted our analysis on the 1,054 counties or equivalents that included 20 or more deaths due to CVD per year for all 12 years of our panel. Using an average of 2013 and 2014 population estimates from SEER, these counties represent 87.9% of the US population during our study period, at the midpoint of our panel. Figure 1a. depicts the states that adopted PSL over the 12 years of our panel and the year the policy was adopted. As can be seen in this figure, during this time period, PSL was mostly implemented in the Western and Northeastern US, with only one southern state and Washington DC, implementing this policy in the South and no states implementing this policy in the Midwest. Figure 1b. shows the change in CVD mortality among persons 15-64 in counties included in any of our analyses over the duration of our panel time period.

**Figure 1a.**
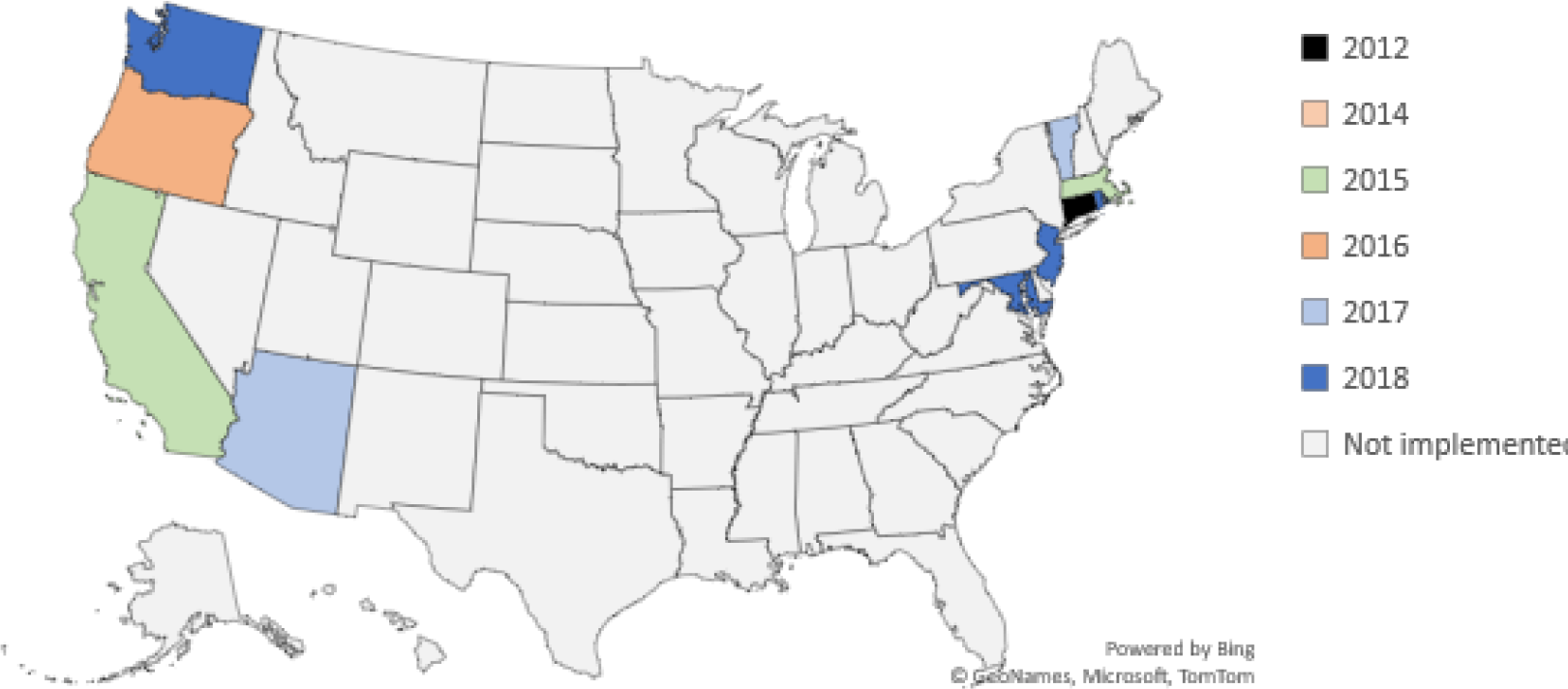
State Level Paid Sick Leave and the Year it Took Effect in the United States, 2008-2019 Source: National Conference of State Legislatures^20^

**Figure 1b.**
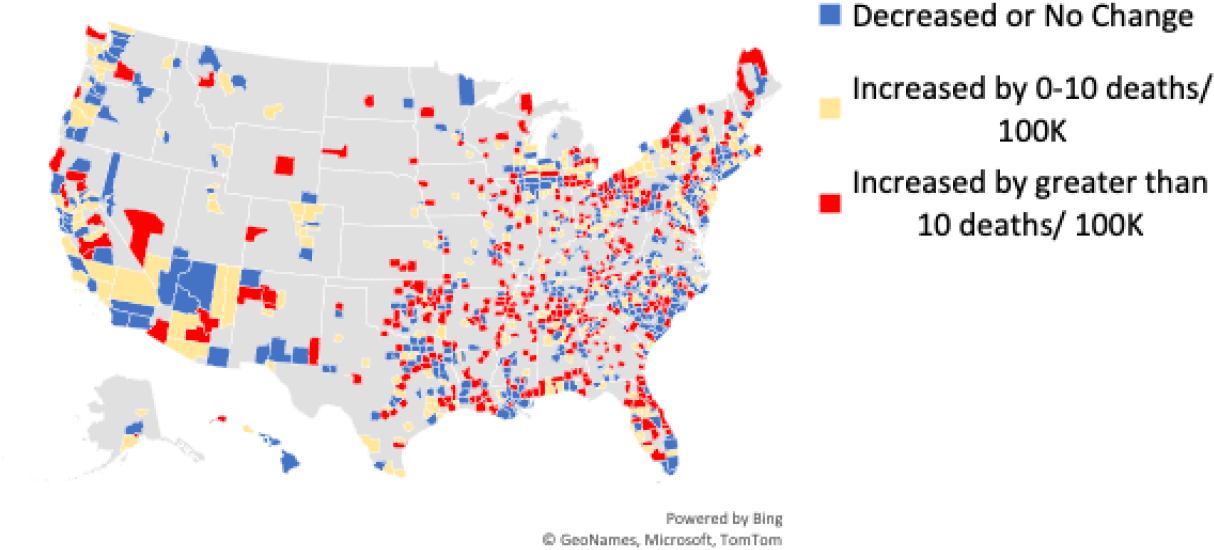
County Level Change in CVD Mortality Rate per 100K Among Persons Age 15-64, 2008 to 2019 (n= 1,054 Counties) Source: CDC Wonder^16^

In Table 1, we display the distribution of time-variant and time-invariant covariates across counties in PSL-adopting states and counties in non-PSL-adopting states. Compared with counties in states that did not adopt PSL policies, counties in the 11 states that adopted PSL had a significantly higher baseline (2008) median income and a higher change in median income over the panel time period, on average. Counties in PSL states had a significantly lower mean percent of persons living in poverty at baseline than counties in non-PSL states. However, there were no differences in the mean change in poverty rates between these two groups throughout the panel. Counties in PSL states had a significantly higher mean baseline unemployment rate than counties in non-PSL states and a higher mean reduction in the unemployment rate over the duration of the panel. Finally, in counties in PSL states, we found that there was both a significantly lower average rate of persons without health insurance at baseline and a significantly larger average reduction in the rate of persons without health insurance throughout the panel in comparison with counties in non-PSL adopting states.

**Table 1.**
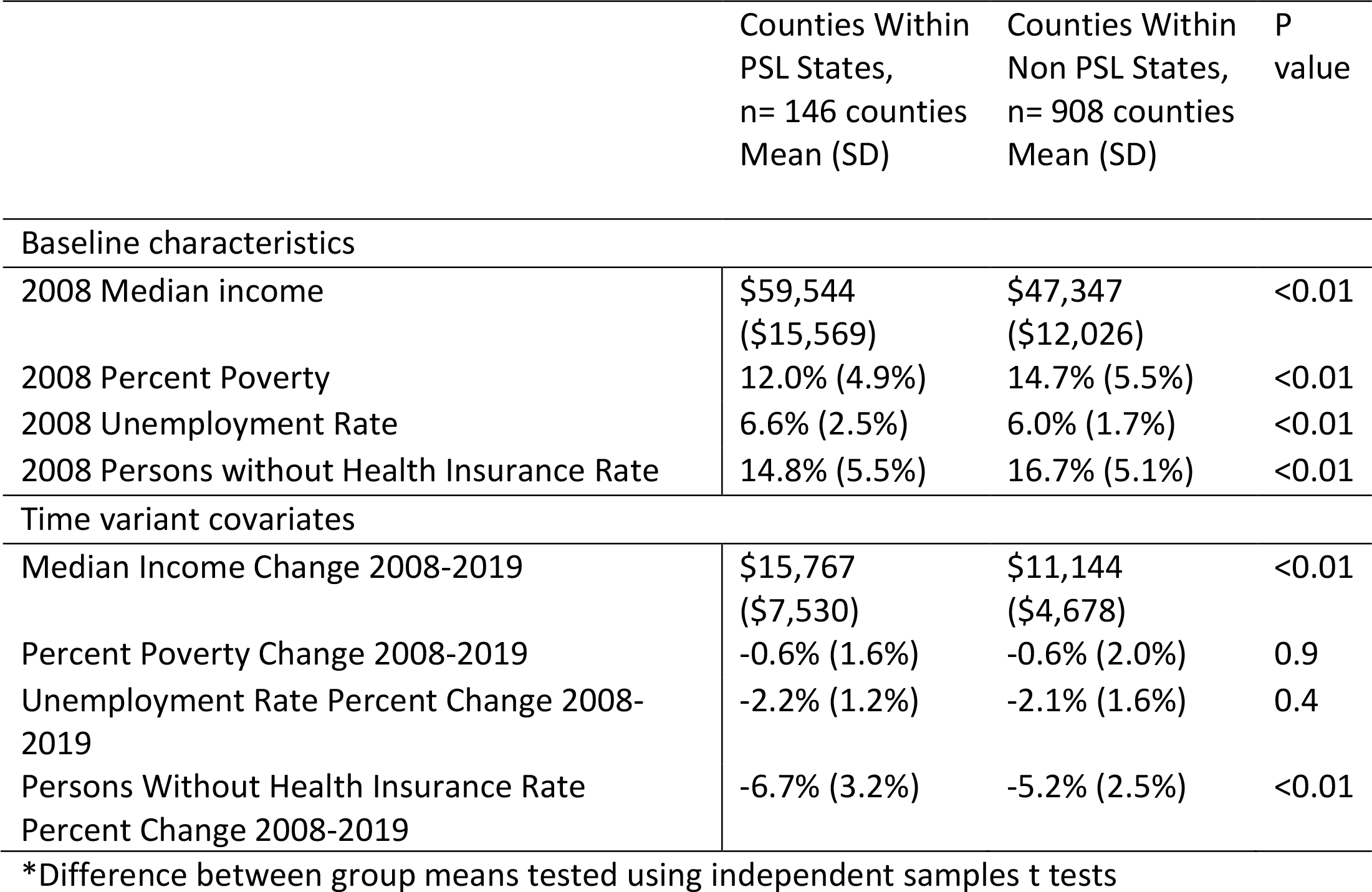
Sociodemographic characteristics of within PSL adopting and non PSL adopting States, United States, 2008-2019.

Figure 2 graphically depicts the results of descriptive analysis plotting the change in county-level CVD mortality rate among persons in counties included in this analysis in PSL-adopting states vs. counties in non-PSL-adopting states within the same region. This figure shows that, in general, in counties in the Northeast, the rate of CVD mortality among persons aged 15 to 64 is lower among counties in states that adopted PSL than counties in non-adopting states, both before and after the years of policy adoption. However, this was not the case in the Western US. In the Western US, the pre-policy and post-policy trends in CVD mortality overlapped, except for Arizona, where the pre- and post-policy trends in CVD mortality were higher in Arizona than the untreated states within the West. Although here we show plots for the South and for counties within the two adopting states, lines look fairly parallel with counties in non-adopting states. These are not included in the main analyses.

**Figure 2.**
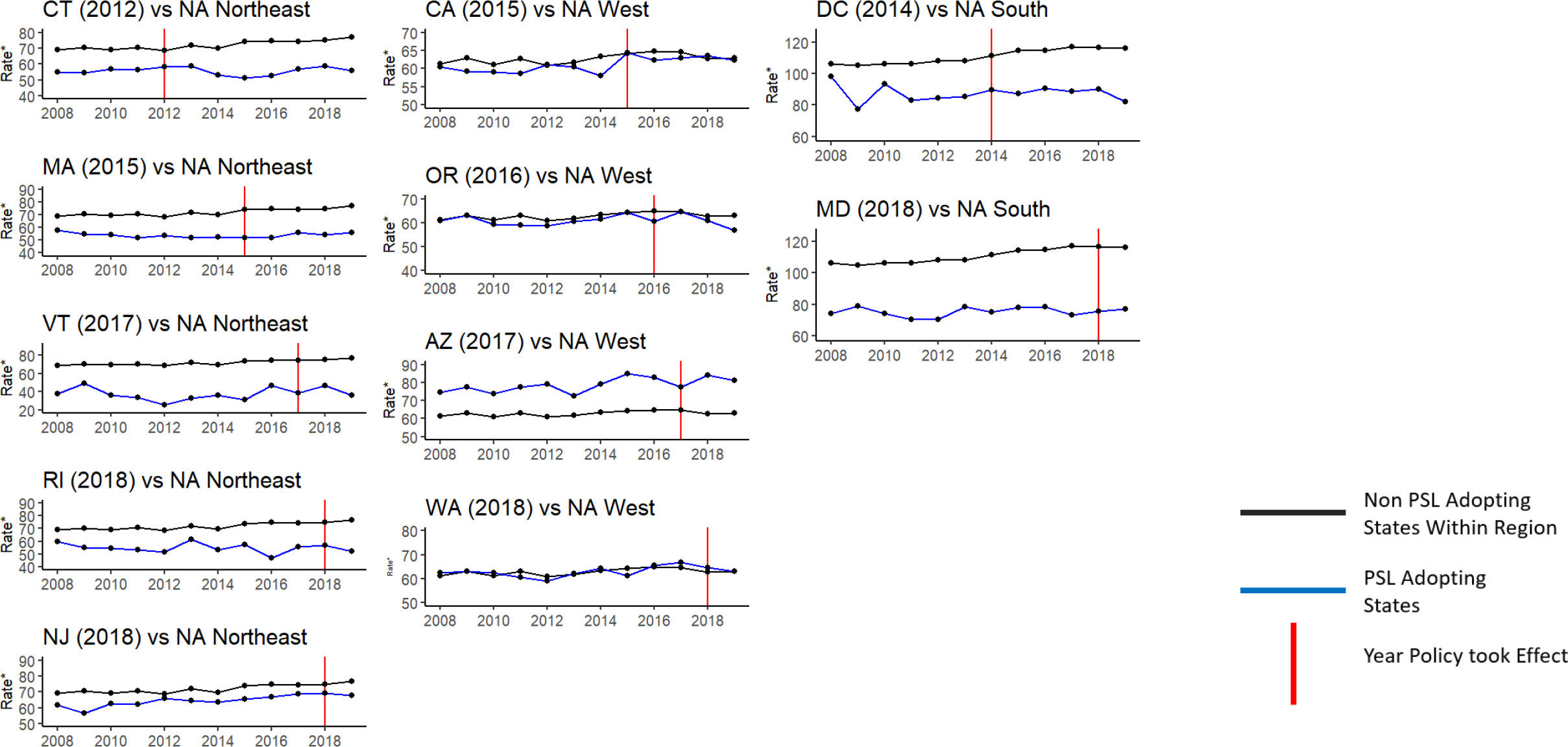
Annual cardiovascular disease mortality rate among persons 15-64* in states that adopted State-level paid sick leave policies (treated) vs non-adopting (NA) states (untreated), 2008 to 2019 *Rates are county-level mortality rates due to cardiovascular disease among persons 15-64, per 100,000 persons. Non-adopting states (as of 2019) in the Northeast are Maine, New Hampshire, Pennsylvania, and New York. Non-adopting states (as of 2019) in West are Colorado, Idaho, New Mexico, Montana, Utah, Nevada, Wyoming, Alaska, and Hawaii. Non-adopting states (as of 2019) in the South are Delaware, Florida, Georgia, North Carolina, South Carolina, Virginia, West Virginia, Alabama, Kentucky, Mississippi, Tennessee, Arkansas, Louisiana, Oklahoma, and Texas

Figure 3 shows the main results of the analysis, focusing on the Northeast and Western regions. Each point is a coefficient from the event study models, representing the difference in the county-level CVD mortality rate ages 15 to 64 between counties in PSL states and non-PSL states that is predicted by the implementation of the PSL policy within a given year. Adjusting for time-variant county-level median income, poverty rate, unemployment rate, and percent of a county without health insurance, the implementation of a PSL policy did not account for differences in county-level CVD mortality rates ages 15 to 64 before the implementation of the policy, with the exception of eight years before treatment.

**Figure 3.**
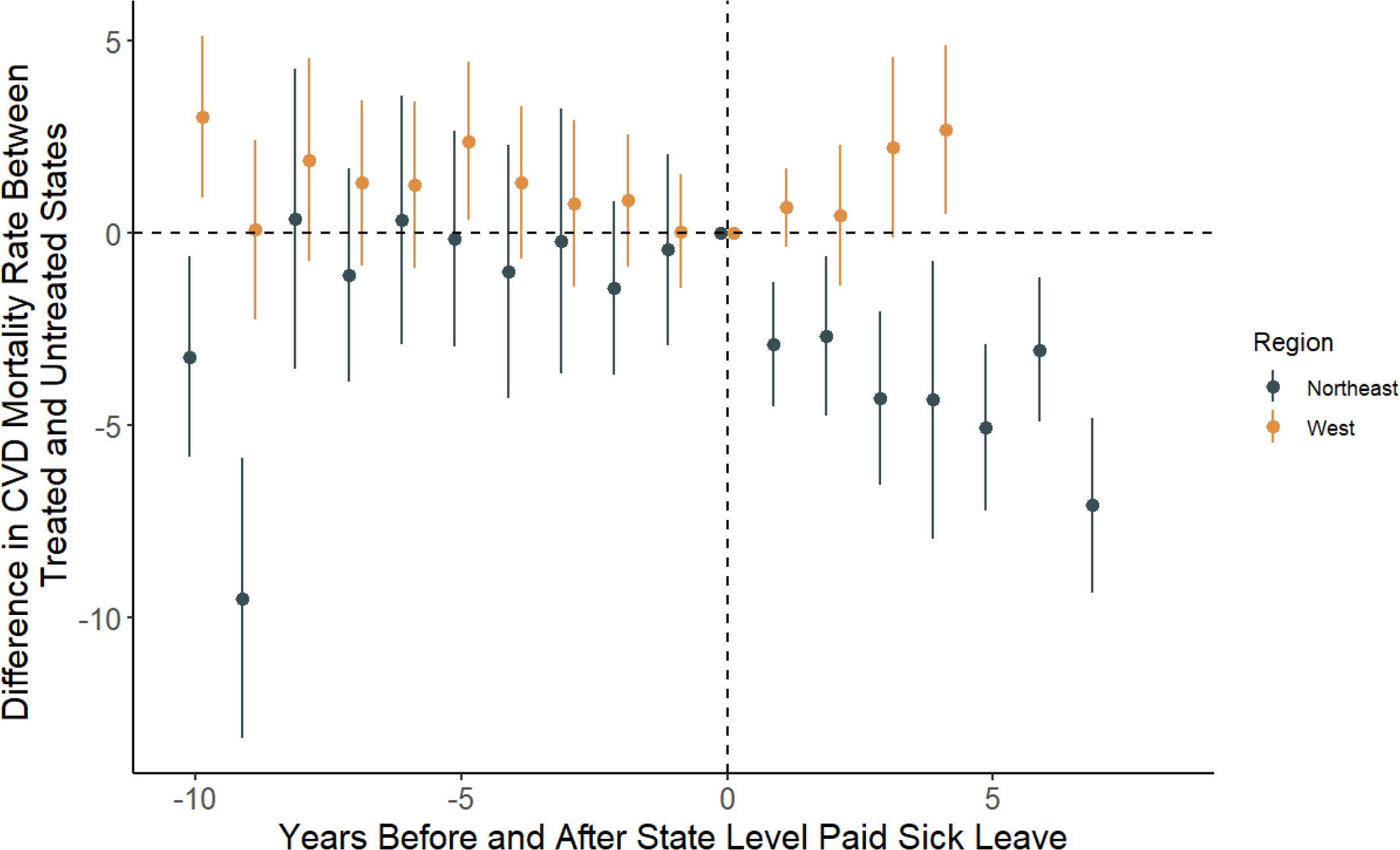
Event study coefficients for change in rates of county-level CVD before and after implementation of paid sick leave, treated states in the Northeast and West compared to untreated states within the same region, 2008-2019 *Models adjusted for time variant county-level median income, poverty rate, unemployment rate, and percent of county lacking health insurance. Mortality rates per 100,000 persons. Adopting States in the Northeast are Connecticut, Massachusetts, Vermont, New Jersey and Rhode Island. Non-adopting states in the Northeast are Maine, New Hampshire, Pennsylvania, and New York. Adopting states in the West are California, Arizona, Oregon and Washington. Nonadopting
states in West are Colorado, Idaho, New Mexico, Montana, Utah, Nevada, Wyoming, Alaska, and Hawaii.

The lack of pretreatment effects, which can be thought of as placebo tests for the intervention, gives us confidence that the post-treatment effects are due to the intervention. In this analysis, we saw a strong pattern for the Northeastern region, in which the coefficients were significant in all years post-treatment, ranging from 7.1 fewer deaths per 100,000 persons (β=-7.1, 95% CI = -9.7, -4.4) seven years post-treatment to 2.7 fewer deaths two years post-treatment (β=-2.7, 95% CI= -5.1, -0.3). This pattern did not hold in the Western region, where the coefficients were all null except for four years after treatment, which was positive. Due to the timing of the implementation of the policies, the counties in states in the Western region did not have follow-up time of more than four years post-treatment. This pattern was similar in models without these covariates (not shown).

In our supplementary material, we include figures for our three sensitivity analyses. Our adjusted models stratified by sex-specific county-level CVD mortality rate age 15 to 64 showed a similar pattern for both the county-level female and male rates, suggesting there is no effect modification by sex. In our sensitivity analysis estimating event study models using age-adjusted rates, we observed a similar pattern to that seen in our main analysis. Finally, in our third sensitivity analysis comparing counties in treated states nationwide to counties in untreated states nationwide, we only saw effects from five through seven years post-treatment.

## DISCUSSION

Within the Northeastern region of the US, we find a strong relationship between state-level implementation of a PSL policy and subsequent reduction in CVD mortality, where five states (CT, MA, VT, RI, and NJ) implemented PSL policies during the panel. In counties in these states, we saw reductions in CVD mortality every year post-policy, resulting in approximately 2.7 to 7.1 fewer deaths per 100,000 people per year in these counties. As counties in this region had CVD mortality rates between 20 and 80 deaths per 100,000 persons per year *before* the policy was implemented, these numbers represent significant drops in the county-level CVD mortality rate. Unlike this beneficial effect of PSL in the Northeastern region of the US, there was no effect of PSL on CVD mortality for states in the Western region.

There are two potential explanations for our findings, 1) that more panel time may be needed to see if the treatment effect is delayed in Western states (where PSL was implemented later), or 2) that there is heterogeneity in the relationship between PSL and county-level CVD mortality for the Eastern United States vs the Western United States. Given the results shown here, there may be heterogeneity in the effect of treatment. The review of these policies by Pomeranz and colleagues details considerable heterogeneity in the number of features of one of these policies, including whether a policy includes days off for the care of a spouse, care of a child, and many other policy characteristics^15^. Importantly, the PSL law in some states included different types of preemption clauses, which may override local policies related to PSL^15^. One study found that state laws that preempt local jurisdictions from changing

PSL laws were associated with suicide and homicide mortality in men^28^. While preemption laws may be an issue, research on this topic from a similar time period showed that employees in the West and Northeast (where the state-level policies were implemented) gained more PSL during this time than persons living in other regions^29^, suggesting that state-level policy is associated with people actually gaining PSL within a state. It is also important to note the pre-policy differences in county-level CVD mortality rates between PSL and non-adopting states in the West are not like those in the Northeast, as California, Oregon, and Washington overlap, and the pre-policy CVD county-level mortality rates in Arizona are greater than that in the non-adopting states. We recommend further investigation on the reasons for differences between the Northeast and West found in this analysis.

Our three sensitivity analyses support the results of our main analysis. There does not appear to be a different pattern when county-level rates of CVD mortality are stratified by sex, which suggests that PSL may be beneficial for both men and women. When we age-adjusted our outcome within the same age bracket, we do not see a change in the patterns of our main analysis, suggesting that differing age distributions (with the bracket of 15 to 64) between the West and Northeast are not accounting for the results of our main analysis. In our analysis comparing counties within untreated states to counties within treated states nationwide, we find significant reductions in county-level CVD mortality rates five, six, and seven years after a policy was implemented. While this result may suggest that the policy could be effective nationwide for non-implementing states (say in the Midwest, for example), it is important to note that this result is most likely driven by the strong results from the Northeastern United States.

There are several mechanistic pathways through which state-level PSL policies may reduce CVD mortality among working-age persons. PSL is associated with more medical checkups for blood pressure, cholesterol, and fasting blood sugar^30^, so it is possible that by promoting preventative medicine checkups, PSL would improve cardiovascular health and reduce the likelihood of CVD mortality.

Additionally, PSL was associated with increased healthcare access in general in another study of Americans^31^. Not having PSL has also been associated with increased odds of psychological distress in the National Health Interview Survey^32^. Psychological distress is, in turn, associated with an increased risk of cardiovascular disease mortality and other cardiovascular outcomes in a large body of literature^33, 34^.

Our analysis has certain limitations. The first is that covariates were collected through several different survey mechanisms, each with its own sampling designs and potential biases. However, all of these sources are the most complete sources of this information for all counties in the US that we are aware of. While not a limitation but a key consideration, this analysis examined county-level rates, county-level covariates, and not individual risk of CVD mortality. For this reason, we urge caution against committing ecological fallacies when interpreting the results of this analysis. This is a consideration rather than a limitation, as county-level CVD mortality rate among working-age adults is a valuable target for public health planning and for supporting the implementation of this policy. Using the CDC mortality data, we were unable to examine individual types of employment or individual income levels, which we recommend be addressed through further research on other types of samples that include this type of information. Another limitation is that we were only able to examine PSL as a binary variable in this research and were not able to evaluate the number of PSL days gained as an ordinal variable as others have done using individual-level data^35^. Finally, we did not include the years 2020 and later in our panel due to the COVID-19 pandemic. Several states implemented these policies in 2020 and 2021, and that information was not used in this analysis. We did not include those years because inference from that time period would add challenges of competing risk (a large number of persons with comorbid diseases dying from COVID-19) as well as drastic, one-time social interventions such as a large decrease in healthcare utilization^36^ extended unemployment benefits^37^, and federal monetary stimulus^38^. Given all those conditions, it would be difficult to isolate the relationship between state-level PSL policy and county-level CVD mortality in 2020 and later. We recommend that future studies examine these years as time passes to isolate the lasting implications of state-level PSL policies.

Our analysis has notable strengths as well. First, the quasi-experimental design controls for both measured and unmeasured time-invariant confounding within a state over the time period of the panel, allowing us to draw much stronger inferences from the results. We also included four important time-variant covariates (median income, percent of county in poverty, unemployment rate, percent without health insurance). We saw strong results in our main analysis after adjustment for these covariates.

Finally, our results were robust to our sensitivity analyses, strengthening our confidence in the findings we present here.

In conclusion, our analysis provides evidence that PSL policies have the potential to be a useful tool for public health professionals and policymakers to prevent and reduce CVD mortality among working-age adults, especially within the Northeastern United States. As more of these policies have been implemented since the time period of our research, we recommend further research on how these policies may be used to prevent cardiovascular disease mortality in other regions and at different jurisdictional levels in the United States.

## Abbreviations

PSL: Paid Sick Leave
CVD: Cardiovascular disease
NA: Non-adopting

## Data Availability Statement

The data and code for this analysis are available as a supplementary file to this manuscript.

## WORKS CITED

1. Control CfD. Leading Causes of Death: National Center for Health Statistics 2022 [Available from: https://www.cdc.gov/nchs/fastats/leading-causes-of-death.htm.

2. Schultz WM, Kelli HM, Lisko JC, Varghese T, Shen J, Sandesara P, et al. Socioeconomic Status and Cardiovascular Outcomes. Circulation. 2018;137(20):2166–78.

3. Lopez AD, Adair T. Is the long-term decline in cardiovascular-disease mortality in high-income countries over? Evidence from national vital statistics. International Journal of Epidemiology. 2019;48(6):1815–23.

4. Abdalla SM, Yu S, Galea S. Trends in cardiovascular disease prevalence by income level in the United States. JAMA network open. 2020;3(9):e2018150–e.

5. Singh GK, Siahpush M, Azuine RE, Williams SD. Widening Socioeconomic and Racial Disparities in Cardiovascular Disease Mortality in the United States, 1969-2013. Int J MCH AIDS. 2015;3(2):106–18.

6. Li J, Loerbroks A, Bosma H, Angerer P. Work stress and cardiovascular disease: a life course perspective. J Occup Health. 2016;58(2):216–9.

7. Adler NE, Newman K. Socioeconomic disparities in health: pathways and policies. Health affairs. 2002;21(2):60–76.

8. Heymann J, Sprague A. Why Adopting a National Paid Sick Leave Law Is Critical to Health and to Reducing Racial and Socioeconomic Disparities—Long Past Due. JAMA Health Forum. 2021;2(5):e210514–e.

9. Conway SH, Pompeii LA, Roberts RE, Follis JL, Gimeno D. Dose-Response Relation Between Work Hours and Cardiovascular Disease Risk: Findings From the Panel Study of Income Dynamics. J Occup Environ Med. 2016;58(3):221–6.

10. KivimäKi M, Batty GD, Ferrie JE, Kawachi I. Cumulative meta-analysis of job strain and CHD. Epidemiology. 2014;25(3):464–5.

11. Kim D. Paid Sick Leave and Risks of All-Cause and Cause-Specific Mortality among Adult Workers in the USA. Int J Environ Res Public Health. 2017;14(10).

12. Sultan-TaïEb H, Chastang J-F, Mansouri M, Niedhammer I. The annual costs of cardiovascular diseases and mental disorders attributable to job strain in France. BMC Public Health. 2013;13(1):748.

13. Matilla-Santander N, Muntaner C, Kreshpaj B, Gunn V, Jonsson J, Kokkinen L, et al. Trajectories of precarious employment and the risk of myocardial infarction and stroke among middle-aged workers in Sweden: A register-based cohort study. The Lancet Regional Health-Europe. 2022;15:100314.

14. Heymann J, Rho HJ, Schmitt J, Earle A. Ensuring a healthy and productive workforce: comparing the generosity of paid sick day and sick leave policies in 22 countries. International Journal of Health Services. 2010;40(1):1–22.

15. Pomeranz JL, Silver D, Lieff SA, PagáN JA. State Paid Sick Leave and Paid Sick-Leave Preemption Laws Across 50 U.S. States, 2009-2020. Am J Prev Med. 2022;62(5):688–95.

16. National Vital Statistics System, Mortality 1999-2020 on CDC WONDER Online Database, released in 2021 [Internet]. [cited July 25th, 2023].

17. Health NYSDo. Rates Based on Small Numbers-Statistics Teaching Tools New York Department of Health2023 [Available from: https://www.health.ny.gov/diseases/chronic/ratesmall.htm.

18. Workers Under 18. In: Labor USDo, editor. Online2023.

19. Munnell AH. What is the average retirement age. Center for Retirement Research. 2011;11(11):1–7.

20. Legislators NCoS. Paid Sick Leave Coverage and Benefits 2022 [Available from: https://www.ncsl.org/research/labor-and-employment/paid-sick-leave.aspx.

21. MiniñO A, Klein R. Mortality From Major Cardiovascular Diseases: United States, 2007. National Center for Health Statistics. 2010:2020–07.

22. Bell W, Basel W, Cruse C, Dalzell L, Maples J, O’Hara B, et al. Use of ACS data to produce SAIPE model-based estimates of poverty for counties. Census Report. 2007.

23. Current Employment Statistics [Internet]. Available from: https://www.bls.gov/ces/.

24. Bowers L, Gann C, Upton R. Small area health insurance estimates: 2016. Small Area Estimates Current Population Reports. 2018.

25. Corporation M. Microsoft Excel 2018.

26. Census US, cartographer Census Regions of the United States Online: U.S. Census Bureau American Community Survey Office 2019.

27. Team R. RStudio: Integrated Development for R. RStudio. Boston, MA2020.

28. Wolf DA, Montez JK, Monnat SM. U.S. State Preemption Laws and Working-Age Mortality. Am J Prev Med. 2022;63(5):681–8.

29. Johnson CY, Said K, Price AE, Darcey D, ØStbye T. Paid sick leave among US private sector employees. Am J Ind Med. 2022;65(9):743–8.

30. Derigne L, Stoddard-Dare P, Collins C, Quinn L. Paid sick leave and preventive health care service use among U.S. working adults. Prev Med. 2017;99:58–62.

31. Hegland TA, Berdahl TA. High Job Flexibility And Paid Sick Leave Increase Health Care Access And Use Among US Workers. Health affairs. 2022;41(6):873–82.

32. Stoddard-Dare P, Derigne L, Collins CC, Quinn LM, Fuller K. Paid sick leave and psychological distress: An analysis of US workers. American Journal of Orthopsychiatry. 2018;88(1):1.

33. Dimsdale JE. Psychological stress and cardiovascular disease. Journal of the American College of Cardiology. 2008;51(13):1237–46.

34. Mcguire AW, Ahearn E, Doering LV. Psychological distress and cardiovascular disease. Journal of Clinical Outcomes Management. 2015;22(9):421–32.

35. Derigne L, Stoddard-Dare P, Quinn LM, Collins C. How Many Paid Sick Days Are Enough? J Occup Environ Med. 2018;60(6):481–9.

36. Moynihan R, Sanders S, Michaleff ZA, Scott AM, Clark J, To EJ, et al. Impact of COVID-19 pandemic on utilisation of healthcare services: a systematic review. BMJ open. 2021;11(3):e045343.

37. Acs G, Karpman M. Employment, income, and unemployment insurance during the Covid-19 Pandemic. Urban Institute. 2020:1–11.

38. Chu L, Teng L. Does Stimulus Check Payment Improve People’s Mental Health in the COVID-19 Pandemic? Evidence from US Household Pulse Survey. The Journal of Mental Health Policy and Economics. 2022;25(4):133–42.

